# Strong reduction of distal colorectal cancer incidence and mortality but no reduction of proximal colon cancer after screening colonoscopy: prospective cohort study

**DOI:** 10.1101/2020.03.26.20044115

**Authors:** Feng Guo, Chen Chen, Bernd Holleczek, Ben Schöttker, Michael Hoffmeister, Hermann Brenner

## Abstract

**Background:** A claimed advantage of colonoscopy over sigmoidoscopy in colorectal cancer (CRC) screening is prevention of CRC not only in the distal colon and rectum but also in the proximal colon. We aimed to assess the association of screening colonoscopy use with overall and site-specific CRC incidence and associated mortality.

**Methods:** Information on use of screening colonoscopy as well as potential confounding factors was obtained at baseline in 2000-2002, updated at 2-, 5-, and 8-year follow-up from 9207 participants aged 50-75 years without history of CRC in a statewide cohort study in Saarland, Germany. Covariate-adjusted associations of screening colonoscopy with CRC incidence and mortality, which were obtained through record linkage with the Saarland Cancer Registry and mortality statistics up to 2016, were assessed by Cox proportional hazards models with time-varying exposure information.

**Findings:** During a median follow-up of 15·3 years, 227 participants were diagnosed with CRC and 81 died from CRC. Screening colonoscopy was associated with strongly reduced overall CRC incidence (adjusted hazard ratio, aHR 0·54, 95% confidence interval, CI 0·41-0·72) and mortality (aHR 0·39, 95% CI 0·24-0·63). However, strong incidence and mortality reduction was seen for distal CRC (aHRs 0·44, 95% CI 0·30-0·63, and 0·35, 95% CI 0·19-0·66, respectively) only, but not for proximal CRC (aHRs 0·99, 95% CI 0·58-1·68, and 0·72, 95% CI 0·29-1·81, respectively).

**Conclusion:** In this large prospective cohort study from Germany, screening colonoscopy was associated with strong reduction in total and distal CRC incidence and mortality, but no reduction was seen for cancer incidence and mortality in the proximal colon.

**Research in context:** *Evidence before this study:*

- Multiple randomized controlled trials have demonstrated that screening with flexible sigmoidoscopy can substantially reduce incidence and mortality from cancer in the distal colon and rectum.
- Evidence on the impact of screening colonoscopy on colorectal cancer incidence and mortality from randomized trials is lacking, and evidence from prospective cohort studies is very limited.
- In particular, it is highly uncertain to what extent screening colonoscopy can additionally reduce incidence and mortality from cancer in the proximal colon.

*Added value of this study:*

- This population-based, prospective statewide cohort study from Saarland/Germany with repeat updates of exposure information demonstrates major reduction of total and distal CRC incidence and mortality among people who underwent screening colonoscopy.
- However, no reduction of incidence and mortality from cancer in the proximal colon was observed.
- These results challenge the expectation of incremental effectiveness of colonoscopy screening over screening by flexible sigmoidoscopy in preventing colorectal cancer.

*Implications of all the available evidence:*

- Our results may impact on recommendations, offers and use of colonoscopy versus flexible sigmoidoscopy for colorectal cancer screening.

## Background

Colorectal cancer (CRC) is the third most common cancer globally, accounting for more than 1·8 million new cancer diagnoses per year.(1) Most CRCs slowly develop over many years from adenomas which can be detected and removed by endoscopic screening. The two main options for endoscopic screening are flexible sigmoidoscopy (FS), which enables visualization of the distal colon and rectum, where the majority of CRCs are located, and colonoscopy, which enables visualization of the entire colon and rectum. However, the more complete visualization by screening colonoscopy comes at the prize of the need of complete bowel cleansing, a major obstacle to screening adherence, substantially higher costs, and higher complication rates. A crucial question for choosing between colonoscopy and FS for CRC screening is therefore if and to what extent this prize is justified by colonoscopies’ ability to additionally visualize the proximal colon and to prevent cancers in the proximal colon.

Multiple randomized controlled trials (RCTs) have demonstrated reduction of distal CRC incidence and mortality by FS-based screening.(2-5) RCT results on long-term effects of screening colonoscopy will become available only many years from now,(6) and evidence from prospective cohort studies keeps being very limited. A number of case-control studies have consistently suggested that screening colonoscopy was associated with strongly reduced CRC incidence and mortality,(7-12) and they typically found stronger protection from cancer in the distal colon and rectum than from proximal colon cancer.(13) However, apart from availability of limited covariate data in some of the studies, case-control studies on screening are prone to specific potential biases.(14) The few prospective cohort studies were either very limited in sample size (15) or follow-up time (16) and measured colonoscopy exposure just once, i.e., at recruitment only,(15,16) or focused on specific professional groups (such as female teachers or health care professionals) or age groups only (17-19) and on CRC incidence (19) or mortality only.(18)

Germany was one of the first countries to offer colonoscopy as a primary CRC screening examination nationwide. Since October 2002, men and women aged 55 years or older have been entitled to have up to two screening colonoscopies 10 or more years apart.(20) We aimed to assess the association between use of screening colonoscopy and CRC incidence and mortality in a prospective population-based cohort study of older adults from Germany, paying particular attention to specific effects of preventing cancer in the proximal and distal colon and rectum.

## Methods

### Study design and study population

Our analysis is based on data from the ESTHER study, an ongoing statewide population-based cohort study among older adults conducted in Saarland, Germany. Details of the study design have been reported elsewhere.(21, 22) Briefly, 9,949 male and female residents of Saarland aged 50-75 years with sufficient knowledge of the German language were recruited in 2000-2002 by their general practitioners (GPs) during a routine health check-up aiming at early detection of cardiovascular diseases and diabetes. The study population has been shown to closely resemble the study population of a representative German national health survey within the corresponding age range carried out in 1998 with respect to major sociodemographic and health related characteristics.(23) ESTHER participants and their GPs are regularly re-contacted every 2 to 3 years, and participants are followed up with respect to incidence and mortality of major chronic diseases including cancer. The study was approved by ethics committees of the Medical Faculty Heidelberg of Heidelberg University and of the Physicians’ board of Saarland. Written informed consent was obtained from each participant.

For the current analysis, we excluded participants with missing information on screening colonoscopy before recruitment (n=742). For analyses on CRC incidence, we additionally excluded participants with a CRC diagnosis before recruitment (n=111), leaving a total of 9096 participants for analysis of CRC incidence and 9207 participants for analysis of CRC mortality.

### Data collection

At baseline, comprehensive information on lifestyle factors and medical history was obtained by self-administered standardized questionnaire from the participants, which was complemented by GP information from the health check-ups and medical records. In particular, participants were asked if they ever had a colonoscopy for screening purposes, and if so, the date of the most recent examination. Follow-up information on screening colonoscopy was obtained by standardized questionnaires at 2-, 5-, and 8-year follow-up, conducted in 2002-2004, 2005-2007, and 2008-2010, respectively. In the few cases where the date of screening colonoscopy was missing (3·8% of screening colonoscopies reported in follow-up questionnaires), the date was set at the midpoint of the respective time interval between consecutive follow-up rounds. If a CRC was diagnosed during that time interval, we assumed that CRC was diagnosed at this screening colonoscopy, and the date of screening colonoscopy was assumed to equal the date of CRC diagnosis.

Follow-up with respect to CRC incidence by the end of 2016 was conducted through record linkage with the statewide Saarland Cancer Registry. Vital status by the end of 2016 and date of death could be ascertained by record linkage with population registries for 99·7% of the cohort. Information on cause of death could be obtained from 98·9% of deceased participants from public health authorities. Hence, analysis of CRC incidence included CRC-free person-times under observation and cases diagnosed with CRC (International Classification of Diseases, 10^th^ Revision [ICD 10] codes C18-C20) and analysis of CRC mortality included person-times and deaths from CRC (ICD 10 codes C18-C20) until the end of 2016.

### Statistical analyses

We first described the study population with respect to sociodemographic characteristics and other known or suspected CRC risk factors, including age, sex, school education (≤9, 10-11, ≥12 years of schooling), history of CRC in a 1^st^ degree relative, smoking (never, former, current), alcohol consumption (women: none, <20, ≥ 20 g/day; men: none, <40, ≥40 g/day), body mass index (<25, 25-29·9, ≥30 kg/m^2^), physical activity (<1, 1-2, ≥2 hours/week of vigorous physical activity), red and processed meat consumption (≤1 time/week, multiple times per week, ≥1 time/day), use of hormone replacement therapy (never, former, current; women only), regular use of aspirin, and physician-diagnosed diabetes.

Exposure status was initially defined at baseline, and updated at 2-, 5-, and 8-year follow-ups. Baseline characteristics of participants who used screening colonoscopy and participants who never had screening colonoscopy before or after study enrollment were compared using chi-square tests. Cox proportional hazards models with screening colonoscopy as time-varying exposure variable (in that participants switched from unexposed to exposed when they had a screening colonoscopy during follow-up) were used to assess associations with CRC incidence and mortality, accounting for the above-mentioned covariates. We included person-time from start of enrollment to CRC incidence, death, or end of 2016 (whatever came first) for analyses of CRC incidence and from start of enrollment to CRC death, death from other cause, or end of 2016 (whatever came first) for analyses of CRC mortality. The proportional hazards assumption was evaluated and accounted for where necessary by interaction terms between covariates and time since enrollment. Associations with CRC incidence and mortality were assessed for ever versus never use of screening colonoscopy and, in addition, for use of screening colonoscopy within the preceding 10 years versus never use.

In all analyses, two types of models were run. Model 1 adjusted for sex and age only, and model 2 adjusted for all of the above-listed covariates that were associated with ever use of screening colonoscopy with a p-value <0·2. Multiple imputation by chained equations (24) was applied to deal with missing values in the following covariates (missing values in parentheses): school education (n=212, 2·3%), history of CRC in a 1^st^ degree relative (n=105, 1·1%), smoking (n=226, 2·5%), alcohol consumption (n=825, 9·0%), body mass index (n=15, 0·2%), physical activity (n=26, 0·3%), red meat consumption (n=475, 5·2%), processed meat consumption (n=429, 4·7%), use of hormone replacement therapy (n=1, 0·02%), diabetes (n=131, 1·4%). The imputation procedure was applied under the assumption of data missing at random and five datasets were imputed using the variables included in the fully adjusted Cox regression models. Associations between screening colonoscopy and CRC incidence and mortality were quantified by hazard ratios and their 95% confidence intervals.

Separate models were run for total, proximal and distal CRC incidence and mortality. CRCs were defined as proximal if they were located proximal of the splenic flexure, and as distal otherwise. Hazard ratios by CRC subsites were estimated using a competing risk method where cases from the complementary site were censored at the date of diagnosis. We used the Wald test to compare hazard ratios between sites (i.e., proximal vs. distal). Sex-specific analyses on the association of screening colonoscopy with total and site-specific CRC incidence were conducted in addition to analyses for the whole study population.

In order to explore net effects of screening colonoscopy on total cancer incidence and mortality and to evaluate potential residual confounding related to health consciousness, we also quantified the association of screening colonoscopy with overall and non-CRC cancer incidence and mortality.

All statistical analyses were performed by the software package SAS (version 9·4; SAS Institute Cary, North Carolina, USA) and R version 3·3·2.(25) Two-sided p-values <0·05 were considered statistically significant.

## Results

Table 1 shows characteristics of the study participants. Mean age at recruitment was 61 years, 54·8% were women. Among those still alive, 96%, 89% and 72% of participants returned follow-up questionnaires at 2-, 5- and 8-year follow-up, respectively. By the 8-year follow-up, 5175 participants (56·2%) had reported to have ever undergone a screening colonoscopy. Approximately 9% of participants had a first-degree relative with CRC, and these participants were more likely to have had a screening colonoscopy (66·7%) than participants without a family history (55·4%, p<0·001). Other factors associated with use of screening colonoscopy were school education, smoking, alcohol consumption, body mass index, physical activity, red and processed meat consumption, use of hormone replacement therapy, and diabetes.

**Table 1.** Characteristics of the ESTHER study population included in the analysis. Missing values for the following items: education (n=212), history of CRC in a 1^st^ degree relative (n=105), smoking status (n=226), alcohol consumption (n=825), body mass index (n=15), physical activity (n=26), red meat consumption (N=475), processed meat consumption (n=429), hormone replacement therapy (n=1), aspirin intake (N=108), and diabetes (n=131).

| Characteristic |  | Overall<br>(N=9207)<br>N (column %) | Ever use of screening<br>colonoscopy up to<br>8-year follow-up<br>N (row %) | p-value* |
| --- | --- | --- | --- | --- |
| Age [years] | <65 | 5744 (62.4) | 3331 (58.0) | <0.001 |
|  | ≥65 | 3463 (37.6) | 1844 (53.2) |  |
| Sex | Male | 4158 (45.2) | 2347 (56.4) | 0.69 |
|  | Female | 5049 (54.8) | 2828 (56.0) |  |
| Education [years] | ≤9 | 6657 (74.0) | 3617 (54.3) | <0.001 |
|  | 10-11 | 1304 (14.5) | 795 (61.0) |  |
|  | ≥12 | 1034 (11.5) | 663 (64.1) |  |
| History of CRC in a 1<sup>st</sup><br>degree relative | No | 8252 (90.7) | 4571 (55.4) | <0.001 |
|  | Yes | 850 (9.3) | 567 (66.7) |  |
| Smoking status | Never | 4474 (49.8) | 2514 (56.2) | <0.001 |
|  | Former | 2986 (33.3) | 1808 (60.5) |  |
|  | Current | 1521 (16.9) | 747 (49.1) |  |
| Alcohol consumption<sup>†</sup> | Abstainer | 2670 (31.8) | 1403 (52.5) | <0.001 |
|  | Intermediate | 5126 (61.2) | 3029 (59.1) |  |
|  | High | 586 (7.0) | 350 (59.7) |  |
| Body mass index [kg/m<sup>2</sup>] | <25 | 2545 (27.7) | 1481 (58.2) | <0.001 |
|  | 25-29.9 | 4337 (47.2) | 2498 (57.6) |  |
|  | ≥30 | 2310 (25.1) | 1188 (51.4) |  |
| Physical activity<sup>-</sup> | Inactive | 1848 (20.1) | 844 (45.7) | <0.001 |
|  | Low | 4214 (45.9) | 2388 (56.7) |  |
|  | Medium or high | 3119 (34.0) | 1932 (61.9) |  |
| Red meat consumption | ≤1 time/week | 3572 (40.9) | 1949 (54.6) | <0.001 |
|  | Multiple times per week | 4568 (52.3) | 2684 (58.8) |  |
|  | ≥1 time/day | 592 (6.8) | 326 (55.1) |  |
| Processed meat<br>consumption | ≤1 time/week | 1728 (19.7) | 1023 (59.2) | 0.02 |
|  | Multiple times per week | 4361 (49.7) | 2488 (57.1) |  |
|  | ≥1 time/day | 2689 (30.6) | 1477 (54.9) |  |
| Hormone replacement<br>therapy<sup>§</sup> | Never | 2563 (50.8) | 1204 (47.0) | <0.001 |
|  | Former | 894 (17.7) | 537 (60.1) |  |
|  | Current | 1591 (31.5) | 1086 (68.3) |  |
| Aspirin intake | No | 8052 (88.5) | 4517 (56.1) | 0.29 |
|  | Yes | 1047 (11.5) | 606 (57.9) |  |
| Diabetes | No | 7672 (84.5) | 4431 (57.8) | <0.001 |
|  | Yes | 1404 (15.5) | 676 (48.1) |  |

CRC colorectal cancer
\* *p*-value for differences in use of screening colonoscopy
† Categories were defined as follows: abstainer, intermediate (women: <20 g/day, men: <40 g/day), high (women: ≥20 g/day, men: ≥40 g/day).
– Defined by hours of vigorous physical activity per week: inactive: <1, low: 1 to <2, medium/high: ≥2
§ among women

During a median follow-up over 15·3 years, 227 incident CRC cases and 81 CRC deaths were observed. Table 2 shows the associations of screening colonoscopy with total, proximal and distal CRC incidence and mortality estimated by Cox proportional hazards models. In age- and sex-adjusted analysis, a history of screening colonoscopy was associated with strongly reduced total CRC incidence and mortality, and these associations were essentially unchanged after controlling for additional potential confounders (adjusted hazard ratio, aHR, 0·54, 95% confidence interval, CI, 0·41-0·72 for incidence and 0·39, 95% CI 0·24-0·63 for mortality). Information on cancer site was available for 205 (90%) of 227 incident cancers and 72 (89%) of 81 CRC-specific deaths. Strong incidence and mortality reduction was seen for distal CRC (aHR 0·44, 95% CI 0·30-0·63 and 0·35, 95% CI 0·19-0·66, respectively), but not for cancer in the proximal colon (aHRs 0·99, 95% CI 0·58-1·68, and 0·72, 95% CI 0·29-1·81, respectively, p-value for heterogeneity in incidence by cancer subsite=0·03). All of the associations were stronger when focusing the analyses on screening colonoscopies conducted within the preceding 10 years, with differences being more pronounced for CRC mortality than for CRC incidence. In particular, screening colonoscopy within the preceding 10 years was also tentatively associated with reduced mortality from proximal cancer, even though the association did not reach statistical significance.

**Table 2.**
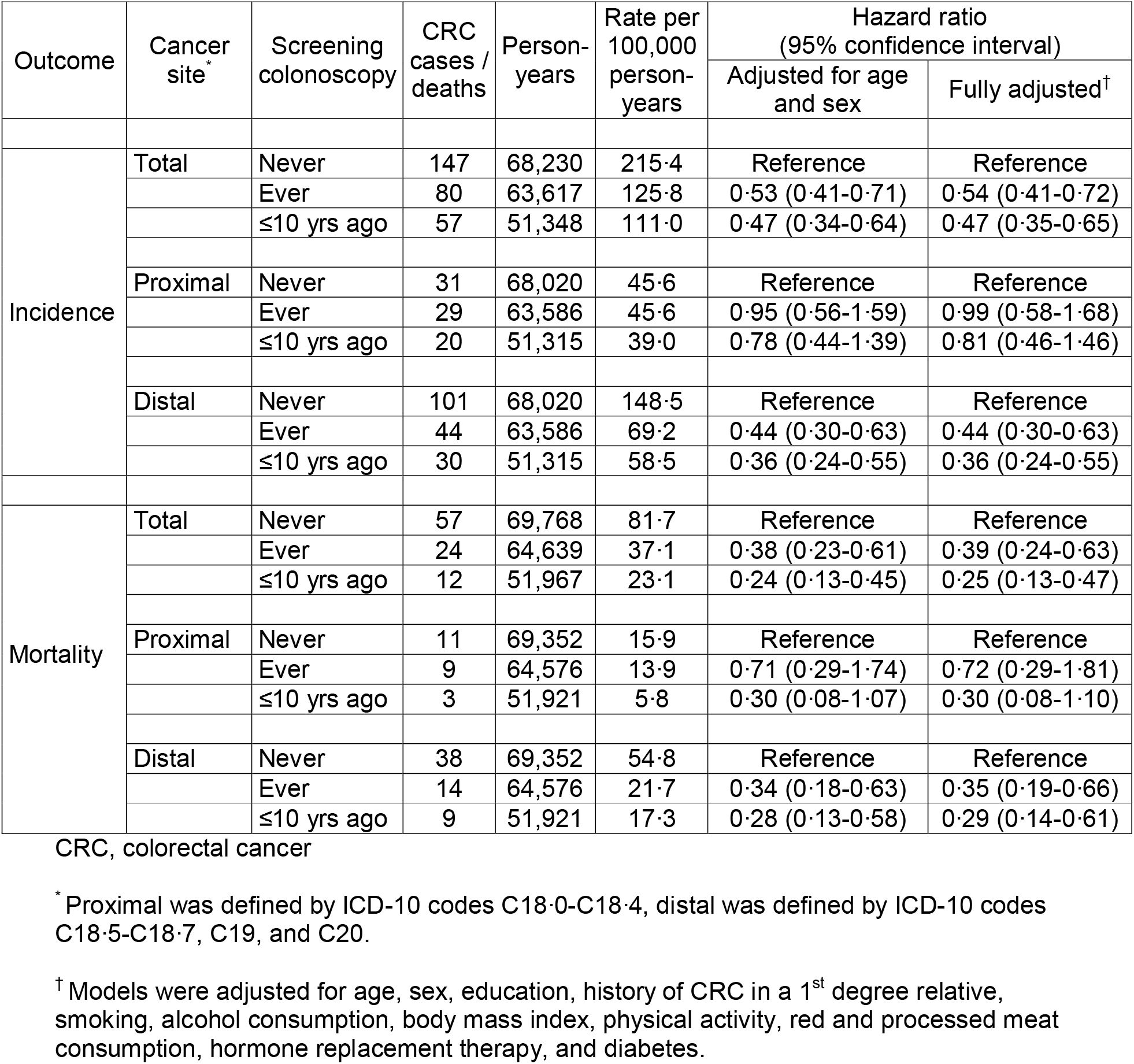
Total and site-specific CRC incidence and mortality according to use of screening colonoscopy

Selective reduction of distal CRC incidence but no reduction of incidence of cancers in the proximal colon was consistently seen among both men and women (Table 3). Associations for distal CRC incidence were tentatively stronger for men than for women but 95% confidence intervals were overlapping.

**Table 3.** Total and site-specific CRC incidence according to sex and use of screening colonoscopy

| Sex | Cancer site | Screening colonoscopy | CRC cases | Person-years | Rate per 100,000 person-years | Hazard ratio (95% confidence interval) |  |
| --- | --- | --- | --- | --- | --- | --- | --- |
|  |  |  |  |  |  | Adjusted for age | Fully adjusted<sup>†</sup> |
| Men | Total | Never | 88 | 29,437 | 298.9 | Reference | Reference |
|  |  | Ever | 46 | 28,058 | 163.9 | 0.51 (0.35-0.73) | 0.51 (0.35-0.74) |
|  |  | ≤10 yrs ago | 33 | 22,693 | 145.4 | 0.44 (0.30-0.67) | 0.45 (0.30-0.67) |
|  | Proximal | Never | 17 | 29,350 | 57.9 | Reference | Reference |
|  |  | Ever | 16 | 28,043 | 57.1 | 0.93 (0.46-1.88) | 0.93 (0.46-1.88) |
|  |  | ≤10 yrs ago | 12 | 22,678 | 52.9 | 0.81 (0.38-1.73) | 0.80 (0.37-1.72) |
|  | Distal | Never | 64 | 29,350 | 218.1 | Reference | Reference |
|  |  | Ever | 26 | 28,043 | 92.7 | 0.40 (0.25-0.63) | 0.39 (0.25-0.63) |
|  |  | ≤10 yrs ago | 17 | 22,678 | 75.0 | 0.32 (0.19-0.55) | 0.32 (0.18-0.55) |
| Women | Total | Never | 59 | 38,793 | 152.1 | Reference | Reference |
|  |  | Ever | 34 | 35,559 | 95.6 | 0.58 (0.38-0.89) | 0.60 (0.39-0.93) |
|  |  | ≤10 yrs ago | 24 | 28,655 | 83.8 | 0.50 (0.31-0.82) | 0.52 (0.32-0.85) |
|  | Proximal | Never | 14 | 38,670 | 36.2 | Reference | Reference |
|  |  | Ever | 13 | 35,543 | 36.6 | 0.97 (0.45-2.09) | 0.99 (0.49-2.39) |
|  |  | ≤10 yrs ago | 8 | 28,637 | 27.9 | 0.74 (0.30-1.78) | 0.82 (0.34-2.03) |
|  | Distal | Never | 37 | 38,670 | 95.7 | Reference | Reference |
|  |  | Ever | 18 | 35,543 | 50.6 | 0.51 (0.28-0.90) | 0.52 (0.29-0.93) |
|  |  | ≤10 yrs ago | 13 | 28,637 | 45.4 | 0.44 (0.23-0.84) | 0.45 (0.23-0.87) |

CRC, colorectal cancer
\* Proximal was defined by ICD-10 codes C18.0-C18.4, distal was defined by ICD-10 codes C18.5-C18.7, C19, and C20.
<sup>†</sup> Models were adjusted for age, education, history of CRC in a 1<sup>st</sup> degree relative, smoking, alcohol consumption, body mass index, physical activity, red and processed meat consumption, hormone replacement therapy, and diabetes.

As can be seen from Table 4, reduction of cancer incidence and mortality was exclusively seen for the distal colon and rectum, with consistent null associations for any other cancer including cancer in the proximal colon. Nevertheless, due to the strong inverse association of screening colonoscopy with distal CRC incidence and mortality, screening colonoscopy was associated with a weak, nonsignificant reduction of overall cancer incidence and a significant reduction of overall cancer mortality (aHRs 0·94, 95% CI 0·85-1·04 and 0·81, 95% CI 0·69-0·94, respectively).

**Table 4.** Associations of screening colonoscopy with incidence and mortality from colorectal cancer and other common cancers

| Cancer site | Incidence |  | Mortality |  |
| --- | --- | --- | --- | --- |
|  | Cases<br>N<sub>Total</sub> | Hazard ratio<br>(95% CI)<sup>*</sup> | Deaths<br>N<sub>Total</sub> | Hazard ratio<br>(95% CI)<sup>*</sup> |
| Colon and Rectum, any subsite | 227 | 0.54 (0.41-0.72) | 81 | 0.39 (0.24-0.63) |
| Proximal colon | 60 | 0.99 (0.58-1.68) | 20 | 0.72 (0.29-1.81) |
| Distal colon or rectum | 145 | 0.44 (0.30-0.63) | 52 | 0.35 (0.19-0.66) |
| Upper gastrointestinal tract<sup>†</sup> | 90 | 1.30 (0.84-2.00) | 49 | 0.87 (0.48-1.55) |
| Pancreas, gallbladder, liver | 124 | 0.81 (0.56-1.16) | 98 | 0.87 (0.58-1.31) |
| Lung | 249 | 1.04 (0.80-1.34) | 184 | 0.88 (0.65-1.18) |
| Breast<sup>-</sup> | 249 | 0.96 (0.74-1.24) | 31 | 0.73 (0.35-1.52) |
| Uterus, ovaries<sup>-</sup> | 63 | 1.05 (0.62-1.77) | 24 | 1.33 (0.57-3.08) |
| Prostate<sup>§</sup> | 309 | 1.07 (0.85-1.35) | 34 | 0.97 (0.48-1.94) |
| Urinary tract | 127 | 1.11 (0.77-1.59) | 33 | 0.50 (0.24-1.11) |
| Hematological malignancy | 121 | 1.02 (0.71-1.48) | 54 | 1.52 (0.85-2.69) |
| Other malignancy | 266 | 1.11 (0.89-1.38) | 71 | 0.81 (0.55-1.20) |
| Any cancer | 1696 | 0.94 (0.85-1.04) | 659 | 0.81 (0.69-0.94) |
| Any cancer other than distal CRC | 1529 | 1.05 (0.95-1.17) | 598 | 0.89 (0.75-1.05) |

CI, confidence interval; CRC, colorectal cancer; N<sub>total</sub>, total numbers of incident cases and deaths included in the analyses, respectively
<sup>\*</sup> Models were adjusted for age, sex, education, history of CRC in a 1<sup>st</sup> degree relative, smoking, alcohol consumption, body mass index, physical activity, red and processed meat consumption, hormone replacement therapy, and diabetes
<sup>†</sup> Including: oral cavity and pharynx, esophagus, stomach
<sup>-</sup> Assessed in female participants only; sex was therefore not adjusted for in Cox model
<sup>§</sup> Assessed in male participants only; sex and hormone replacement therapy were therefore not adjusted for in Cox model

## Discussion

In this prospective population-based cohort of older adults from Germany, incidence and mortality from total and distal CRC was strongly reduced among participants who had undergone screening colonoscopy. However, no reduction was seen in incidence of cancer in the proximal colon, and only a modest, non-significant reduction was seen in mortality from proximal colon cancer.

To our knowledge, this is the first cohort study reporting on the effects of screening colonoscopy on CRC incidence and mortality from Germany, one of the first countries with a nationwide offer of colonoscopy as primary screening examination. Our results are in line with and expand findings of a cross-sectional study among participants of screening colonoscopy in Germany, in which previous colonoscopy was associated with a strongly reduced prevalence of advanced neoplasms in the distal colon and rectum, but not in the proximal colon,(26) and of a population-based case-control study from Germany that also found a strong risk reduction for distal CRC within 10 years after screening colonoscopy.(9) The latter study had also found reduction of risk of cancer in the proximal colon, albeit less pronounced. No results on cancer mortality were available from these studies.

To our knowledge, only one prospective cohort study each has assessed the impact of screening colonoscopy on site-specific CRC incidence and mortality, respectively.(17,18) Our results on site-specific CRC incidence are remarkably consistent with those from a cohort study among female teachers in France with 15 years of follow-up,(17) which reported strong, statistically significant reduction of CRC incidence in the distal colon and rectum (adjusted hazard ratios 0·57 and 0·37, respectively) but not in the proximal colon (adjusted hazard ratio 0·87). A cohort study among female and male health professionals with 22 years of follow-up (18) had also found a protective effect of screening colonoscopy against mortality from cancer in the proximal colon, which was though weaker than the protective effect against distal CRC (adjusted hazard ratios 0·47 and 0·18, respectively).

The estimated reduction of distal CRC incidence and mortality and the lack of reduction of incidence and mortality from proximal CRC among people who underwent screening colonoscopy are also remarkably consistent with estimates of the impact of flexible sigmoidoscopy reported in per-protocol analyses of pertinent RCTs.(13) Taken together, these results support suggestions that colonoscopy strongly protects from CRC overall by detecting and removing precursors of the disease. However, such protection does not seem to apply for cancers in the proximal colon whose precursors are known to differ in many respects from precursors of distal CRCs.

The different effects on proximal and distal CRC incidence and mortality may reflect different routes of carcinogenesis. In particular, proximal cancers more often develop from serrated polyps whose detection and removal pose major challenges to endoscopists.(27, 28) In addition, adenoma miss rates have also been reported to be higher in the proximal colon than in the distal colon and rectum.(28) Notwithstanding lack of protection from incidence of cancers in the proximal colon, colonoscopy may still provide some protection from mortality from cancer in the proximal colon through early detection of preclinical proximal cancer which is expected to result in higher cure rates, given the strong dependency of CRC survival from stage at diagnosis. The point estimate of the hazard ratio for mortality from cancer in the proximal colon in our study within 10 years from screening colonoscopy (0·30) suggests such protection even though this finding failed to reach statistical significance given the small overall number of deaths from proximal CRC.

The selective effects for screening colonoscopy in preventing distal CRC suggested by our study are also in line with and may explain the observation that post-colonoscopy CRCs are much more frequently located in the proximal colon and more often demonstrate microsatellite instability than cancers detected at screening colonoscopy.(29) These patterns support suggestions that different molecular features of proximal cancers may contribute to the lack of effectiveness of colonoscopy in preventing them, besides the more difficult detection of precancerous lesions in the proximal colon.

Furthermore, a previous study has shown an overall sustained improvement in adenoma detection rate from 2003 through 2012 by screening colonoscopy in Germany, which might reflect the favorable development in colonoscopy performance across time periods.(20) Several measures have been considered to improve the quality of German screening colonoscopy program, such as optimized bowel preparation and appropriate withdrawal time (at least 6 minutes).(30, 31) Our findings of absence of protective effects for proximal colon might thus partly reflect poorer quality of earlier colonoscopies.

Like previous studies, our study may underestimate true screening colonoscopy effects (in both the proximal or the distal colon and rectum) to some extent because information on colonoscopies conducted for diagnostic purposes, which are expected to convey similar protection by detecting and removing colorectal neoplasms, was not available. According to national health survey data from Germany, a substantial proportion of older adults also has had diagnostic colonoscopy.(32) This proportion would be expected to be even higher among those not having had screening colonoscopy, as a screening colonoscopy might not be warranted in people who have had a recent diagnostic colonoscopy. The expected “contamination” by diagnostic colonoscopies of the reference group with no screening colonoscopy bears the potential of underestimation of the effects of screening colonoscopy.(33) Screening colonoscopy may therefore have led to somewhat stronger reduction of total and site-specific CRC incidence and mortality than estimated by our study.

The strong effects of screening colonoscopy on total and distal CRC incidence and mortality estimated in this study are in agreement with recent trends of a major decrease in CRC incidence and mortality observed in Germany after nationwide introduction of the offer of screening colonoscopy in 2002.(34) Like in the US, where widespread use of screening colonoscopy already started in the 1990s and an even stronger decline in CRC incidence and mortality has been observed since then,(35) this decline is exclusively seen at older ages covered by CRC screening.(34, 36) However, CRC screening still remains underused, and even stronger and more rapid declines of CRC incidence and mortality could be achieved by enhanced adherence to screening offers,(37) especially among the high risk groups with unfavorable risk factor profiles.

A major reason for non-adherence with offers of screening colonoscopy is the invasive nature of this exam, requiring complete bowel cleansing. Furthermore, costs, complication rates and capacities needed are substantially higher compared to flexible sigmoidoscopy. The findings of a lack of or limited use of screening colonoscopy for reducing cancer incidence in the proximal colon support suggestions that even larger effects might be achieved by offers of flexible sigmoidoscopy, which might be used by larger proportions of the population, in particular if combined with fecal immunochemical testing (which enables detection of the majority of proximal cancers).(38) Cost-effectiveness analyses have commonly assumed substantially stronger total CRC incidence reduction by 10-yearly colonoscopy compared to 5-yearly flexible sigmoidoscopy.(39) Our results suggest that the advantage of screening colonoscopy over flexible sigmoidoscopy may have been overestimated. These suggestions are in line with those of a recent modelling study which suggested similarly strong effects of a single flexible sigmoidoscopy or a single colonoscopy in reducing CRC incidence and mortality.(40) These findings may therefore have important implications for refining analyses of effectiveness and cost-effectiveness of various screening offers and for further development and implementation of screening programs, in particular in countries with limited colonoscopy capacities.

In the interpretation of our study, a number of strengths and limitations deserve careful consideration. Strengths include long-term follow-up of a large cohort from the general population. Repeated follow-up examinations enabled exposure updates every 2 to 3 years, and comprehensive data collection enabled careful adjustment for relevant potential confounders. A major limitation is the lack of information on diagnostic colonoscopies. Also, no update on screening colonoscopies conducted after the 8-year follow-up was available which may have led to some misclassification of exposure and most likely some underestimation of screening colonoscopy effects. Furthermore, despite the overall large size of the cohort, numbers of CRC cases and deaths were still rather small, leading to wide confidence intervals for some of the risk estimates and hindering further analyses for specific high risk subgroups, such as people with a family history of CRC. Although we carefully controlled for multiple potential confounders, we cannot rule out residual confounding by unmeasured confounders. However, the selectivity of effects seen for total and distal CRC and not for any other cancer suggests potential residual confounding (e.g. by factors related to general health consciousness), if any, to be small. To our knowledge, no previous observational study has included such a “selectivity check” of associations.

Despite its limitations, this prospective population-based cohort study adds important evidence on the effects of screening colonoscopy in reducing overall and site-specific CRC incidence and mortality, as well as total cancer mortality. Our results underline the large potential of screening colonoscopy to prevent cancer in the distal colon and rectum and to reduce mortality from these cancers and even total cancer mortality. At the same time, however, our results call for critical re-evaluation of the commonly assumed advantages in effectiveness of screening colonoscopy over screening by flexible sigmoidoscopy. Our results may thereby provide important evidence for refining comparative effectiveness and cost-effectiveness analyses of various screening options and for planning and designing screening programs, in particular in countries with limited colonoscopy resources. They also underline the need of further efforts towards more effective prevention of cancer in the proximal colon.

## Data Availability

Anonymized data relevant to the study may be provided upon reasonable request for uses that are compatible with participants’ informed consent.

## Contributors

HB designed, led and supervised the study and drafted and revised the article. FG conducted the statistical analyses and revised the article. CC contributed to the statistical analyses. BS and BH contributed to the coordination and conduction of data collection and work-up of data. All authors contributed to the interpretation of the data, critically reviewed manuscript drafts, constructively contributed to their finalization and agreed with the final version submitted. The researchers are independent from funders. All authors had full access to all of the data (including statistical reports and tables) in the study and can take responsibility for the integrity of the data and the accuracy of the data analysis.

### Ethical approval

The study was approved by ethics committees of the Medical Faculty Heidelberg of Heidelberg University (ID S-058/2000) and of the Physicians’ board of Saarland.

### Participant consent

Written informed consent was obtained from each participant.

## Acknowledgements

The authors gratefully acknowledge valuable contributions to the conduction of the ESTHER study by Hartwig Ziegler, Christa Stegmaier, Sonja Wolf, Gregor Thal, Martina Mohr, Jenny Peter, Volker Herrmann and Utz Benscheid. The ESTHER study was funded by the Baden-Württemberg State Ministry of Science, Research and Arts (Stuttgart, Germany), the German Federal Ministry of Education and Research (Berlin, Germany), the German Federal Ministry of Family, Senior Citizens, Women and Youth (Berlin, Germany) and the Saarland State Ministry of Social Affairs, Health, Women and Family (Saarbrücken, Germany). The analyses for this project were supported by a grant from the German Cancer Aid (No. 70112095). The funders had no role in the study design; in the collection, analysis, and interpretation of data; in the writing of the report; and in the decision to submit the article for publication. The researchers are independent from funders and all authors had full access to all of the data (including statistical reports and tables) in the study and can take responsibility for the integrity of the data and the accuracy of the data analysis.

